# Long-term transmission chains fueling the drug-resistant tuberculosis burden in Southern Brazil

**DOI:** 10.64898/2026.11.17.26368293

**Authors:** Tainá dos Santos Soares, Andressa Fernandes Pivato, Fernando Frosi, Daniel Brisotto Pavanelo, Fernanda Godinho, Simone Minghelli, Simone de David, Graziele Lima Bello, Katharine Walter, João Perdigão, Ivy Ramis, Pedro Eduardo Almeida da Silva, Richard Steiner Salvato

## Abstract

**Background:** Tuberculosis (TB) remains a leading infectious cause of death, with drug-resistant TB (DR-TB) challenging disease control. Brazil has a high TB burden, and Rio Grande do Sul state reports persistently high incidence and drug-resistance rates. The drivers shaping the regional DR-TB burden remain poorly characterised.

**Methods:** We conducted genomic surveillance of confirmed DR-TB cases in Rio Grande do Sul, Brazil, from January 2023 to September 2024. Clinical isolates underwent genotypic (gDST) and phenotypic drug susceptibility testing (pDST) and whole-genome sequencing (WGS). We analyzed 33 newly sequenced genomes and 267 historical *Mycobacterium tuberculosis* genomes collected in 2011–2019 to characterize resistance, transmission, and spatiotemporal patterns and estimate the contributions of transmitted and acquired rifampicin resistance.

**Findings:** Molecular testing detected rifampicin resistance missed by pDST, including borderline *rpoB* mutations in 18.2% and emerging resistance-associated variants in 3% of the 33 analyzed isolates. Conversely, pDST detected baseline bedaquiline resistance missed by WGS in one newly diagnosed case. Among 300 genomes, 181 (60.3%) clustered into 32 genomic clusters at a 5-SNP threshold, including 12 with long-term circulation and several spanning prisons and communities. Sublineage frequencies remained stable over time, whereas we observed marked geographic structuring and localized clonal expansion. Among 178 rifampicin-resistant isolates, transmission accounted for an estimated 79.2–88.2% of resistance, corresponding to 11.8–20.8% of cases arising through *de novo* acquisition during treatment.

**Interpretation:** Our findings identify active transmission as the main driver of rifampicin-resistant TB in Rio Grande do Sul, with long-term transmission chains extending across both incarcerated and community settings. These results underscore the urgent need to strengthen timely TB case detection and resistance testing, enhance contact investigations, and implement targeted interventions aiming to interrupt persistent transmission chains.

## INTRODUCTION

Tuberculosis (TB) is the leading cause of death from a single infectious agent worldwide, with an estimated 10.6 million new cases and 1.3 million deaths in 2025. Brazil is among the 30 countries with the highest TB burden worldwide, with an estimated incidence of 39.5 cases per 100,000 population^1^. In Brazil, Rio Grande do Sul is a particularly important setting for TB prevention and care. Located at the country’s southernmost border, the state reported a TB incidence of 51.1 cases per 100,000 population in 2025 and over 1,000 new drug-resistant TB (DR-TB) cases between 2015 and 2025^2^.

Drug resistance is one of the greatest challenges to global TB prevention and care, compromising treatment effectiveness, prolonging infectiousness, and increasing mortality and healthcare costs. The emergence of multidrug-resistant (MDR-TB) and extensively drug-resistant TB (XDR-TB) has further complicated care, particularly where access to rapid diagnostics remains limited^3^. DR-TB arises through two pathways: transmission of already resistant strains, or *de novo* acquisition of resistance-conferring mutations during treatment, driven by inadequate regimens, poor adherence, or subtherapeutic drug exposure^4^. Distinguishing between them is essential, because transmitted resistance requires active case-finding and infection prevention and control to interrupt transmission chains, whereas acquired resistance requires effective regimens, adequate drug exposure, and patient-centered adherence support.

Since 2019, World Health Organization (WHO) has prioritized shorter, all-oral regimens including bedaquiline, delamanid, pretomanid, and linezolid^5^. In Brazil, bedaquiline and delamanid were incorporated into the Unified Health System (SUS) in 2020 and became available for routine use in 2021^6^. Nevertheless, data on resistance to these recently introduced drugs and its molecular mechanisms remain scarce in Brazil, where genomic surveillance of *Mycobacterium tuberculosis* (*M. tuberculosis*) remains underutilized despite its resolution for characterizing resistance mutations, identifying lineages, and distinguishing transmitted from acquired resistance^7^.

We therefore used whole-genome sequencing (WGS), integrated with phenotypic and epidemiological data, to characterize the genomic determinants of resistance among DR-TB isolates circulating in Rio Grande do Sul, southern Brazil, estimate the relative contribution of transmitted versus acquired resistance, and identify major genomic clusters and their geographic distribution across the state.

## MATERIALS AND METHODS

### Study population and drug susceptibility testing

Between January 2023 and September 2024, the Central Public Health Laboratory of Rio Grande do Sul (LACEN-RS), the state reference laboratory for mycobacterial culture and drug susceptibility testing, identified 68 culture-positive *M. tuberculosis* isolates from unique patients resistant to rifampicin and/or isoniazid by at least one genotypic or phenotypic test. Samples were submitted by healthcare facilities across the state following suspicion of drug resistance.

Genotypic drug susceptibility testing (gDST) used the GenoType MTBDRplus VER 2.0 line probe assay (LPA; Bruker Corporation, Billerica, MA, USA) according to the manufacturer’s instructions, after DNA extraction from solid cultures with the GenoLyse® VER 1.0 kit. The assay identifies *rpoB* mutations associated with rifampicin resistance and *katG* and *fabG1–inhA* operon promoter mutations associated with isoniazid resistance. Isolates resistant by the first-line LPA or requiring further characterization were tested with the GenoType MTBDRsl VER 2.0 assay, which identifies *gyrA* and *gyrB* mutations associated with fluoroquinolone resistance and *rrs* and *eis* promoter mutations associated with resistance to second-line injectables. Xpert MTB/RIF Ultra results, routinely performed at healthcare units, were retrieved from the Brazilian Laboratory Environment Management System (GAL).

Phenotypic drug susceptibility testing (pDST) was performed at LACEN-RS with the BACTEC MGIT 960 SIRE Kit (Becton Dickinson Diagnostic Systems, Sparks, MD, USA) for rifampicin (critical concentration, 0.5 mg/L), isoniazid (0.1 mg/L), ethambutol (5.0 mg/L), and streptomycin (1.0 mg/L). Isolates were also referred to the Professor Hélio Fraga Reference Center, the national reference laboratory for DR-TB diagnosis, for pDST against amikacin, levofloxacin, moxifloxacin, and bedaquiline.

Isolates were classified according to WHO definitions: MDR-TB as resistance to at least isoniazid and rifampicin; pre-XDR-TB as MDR/RR-TB (rifampicin-resistant TB) with additional resistance to any fluoroquinolone; and XDR-TB as MDR/RR-TB with fluoroquinolone resistance plus resistance to bedaquiline or linezolid. *rpoB* mutations previously reported to confer low-level rifampicin resistance that may be missed by pDST were classified as borderline mutations^8,9^.

### Whole-genome sequencing

A convenience sample of 36 of the 68 available cultures was selected for WGS, based on sequencing cost constraints and aiming to include different geographic regions of the state. DNA was extracted from solid cultures using the cetyltrimethylammonium bromide (CTAB) method^10^. Libraries were prepared with the Illumina DNA Prep kit (Illumina, San Diego, USA) and sequenced on a MiSeq platform with V3 600-cycle chemistry (2×150 bp).

TB-Profiler (v6.3.0)^11^ was used to assign SNP-based lineages and identify fixed and low-frequency (heteroresistant) resistance-associated mutations, using a minimum allele frequency of 10% and a minimum read depth of 10×, against the WHO Catalogue of Mutations in *M. tuberculosis*^12^. We also implemented an in-house pipeline: Trimmomatic v0.39^13^ for adapter removal and quality trimming^13^, FastQC for read-quality verification, BWA-MEM v0.7.19^14^ for alignment to the *M. tuberculosis* H37Rv reference genome (NC_000962.3), and Qualimap v2.2.1^15^ to confirm a minimum average depth of 30×. SNPs and small indels were called with both SAMtools/BCFtools v1.2.1^16^ and GATK HaplotypeCaller v4.6.1.0^17^, retaining only concordant variants^18^. Sites and isolates with more than 10% missing data were excluded, and PE/PPE genes and resistance-associated loci were masked in the whole-genome SNP alignment. Three genomes below the 30× mean-depth threshold were excluded, leaving 33 sequences for downstream analyses.

### Phylogenetic and clustering analysis

To enable temporal comparisons of transmission dynamics and resistance acquisition, we combined the 33 newly sequenced genomes with 267 historical isolates collected in Rio Grande do Sul between 2011 and 2019, comprising previously published sequences^19,20^ and unpublished data from our group, yielding 300 genomes for phylogenetic reconstruction, phylodynamic inference, and genomic cluster identification.

ModelFinder^21^ selected the TVM+F nucleotide substitution model under the Akaike Information Criterion, and a maximum-likelihood phylogeny was reconstructed with IQ-TREE2^22^, using 1,000 ultrafast bootstrap replicates and SH-aLRT, annotated with *ggtree*. Genomic clusters were defined using a 5-SNP threshold^23,24^, with hierarchical clustering (*hclust*, R package ape). Patient addresses, georeferenced at the neighborhood level, were mapped using *ggplot2, sf*, and *geobr*.

### Relative contribution of transmitted versus acquired rifampicin resistance

Rifampicin resistance was assessed based on the presence of resistance-associated mutations in *rpoB*. Patients with no prior history of TB treatment were classified as having transmitted rifampicin resistance, as the absence of previous drug exposure precludes treatment-mediated selection. For previously treated patients, the origin of resistance was determined using a combined clinical and phylogenetic approach implemented in the *TB-SIMEX* pipeline (https://github.com/RuanSpies21/TB-SIMEX)^25^, mapping resistance mutations onto the phylogeny and applying two complementary approaches (N and N−1) to define the plausible range of resistance origins. Complete parameters and scripts are available in **Supplementary Methods**.

### Temporal, spatial, and diversity patterns of *M. tuberculosis* sublineages

Lineage assignments, collection years, and municipality of origin were analyzed in R (*dplyr, tidyr, ggplot2*). Sublineages represented by fewer than five isolates were excluded from spatial inferential analyses, geographic association tests were restricted to the ten municipalities with the most sampled isolates, and sublineages were aggregated at the third lineage level for temporal analyses. Within-municipality sublineage diversity was quantified using Simpson’s Diversity Index (1 − D), ranging from 0 (complete dominance of a single sublineage) to 1 (high diversity). Fisher’s exact test was used to calculate exact odds ratios (ORs) with 95% confidence intervals (CIs) for the association between dominant sublineages and their respective municipalities. Temporal and geographic structuring of sublineages was evaluated using Pearson’s Chi-square test of independence. Because many cells had expected frequencies below five, p-values were obtained by Monte Carlo simulation with 10,000 permutations. When a significant overall geographic association was detected, post-hoc analysis of standardized residuals identified the municipality–sublineage combinations contributing most strongly to it, with absolute values ≥ 1.96 considered significant (p < 0.05).

### Ethics statement

This study was approved by the Research Ethics Committee of the Escola de Saúde Pública do Rio Grande do Sul (CEP/ESP-RS), Brazil (approval number 56756122.5.0000.5312). Individual informed consent was waived due to the public health surveillance nature of the study and the use of de-identified patient data.

## RESULTS

In 2023 and 2024, Rio Grande do Sul reported 152 new cases of DR-TB. Over the 21-month study period, 68 isolates were referred to LACEN-RS. Of these, 36 were selected for WGS, and 33 passed bioinformatic quality control, the three excluded had mean coverage below 30×. The 33 included isolates did not differ significantly from the 35 not included in patient age, sex, geographic origin, MGIT-based phenotypic profiles, or mutation patterns detected by line probe assays (p > 0.05; **Supplementary Table 1**). Patients came from 14 municipalities, most frequently Porto Alegre (n=10), Canoas (n=7), and Alvorada (n=3). The remaining municipalities contributed one or two isolates each. Patients were predominantly male (78.8%), aged 19 to 71 years (mean 37.2), and seven (21.2%) were incarcerated at the time of diagnosis. Based on pDST and available molecular testing (Xpert MTB/RIF Ultra and LPA), the 33 isolates were classified as 15 MDR, 13 isoniazid-resistant, two pre-XDR-TB, two RR-TB, and one resistant to both isoniazid and bedaquiline.

Isoniazid resistance was detected by pDST in 31/33 isolates (93.9%), of which 26 (83.9%) were also resistant by LPA (**Figure 1A**), mainly through the *katG* MUT1 (Ser315Thr; n = 21) and *inhA* MUT1 (−15C>T; n = 4) probes. Of the five pDST-resistant isolates without LPA-detected mutations, WGS identified *katG* Asp735Ala, outside the LPA target region, in one (a pre-XDR-TB isolate). No resistance-associated mutation was found in the other four or in the two pDST-susceptible isolates. For rifampicin, 11/33 isolates (33.3%) were resistant by pDST (**Figure 1B**), Xpert MTB/RIF Ultra and LPA detected 10 of these (90.9%), mostly through the MUT3 probe (*rpoB* Ser450Leu; n = 6). The only isolate missed by both assays carried a 12-nucleotide *rpoB* insertion associated with a genomic cluster endemic to Rio Grande do Sul^26^. Conversely, 9 of the 22 pDST-susceptible isolates (40.9%) were resistant by at least one molecular method. WGS explained seven of these discordances with borderline *rpoB* mutations (Asp435Tyr, n = 6; and Leu430Pro, n = 1, at 42% of reads, consistent with heteroresistance).

**Figure 1.**
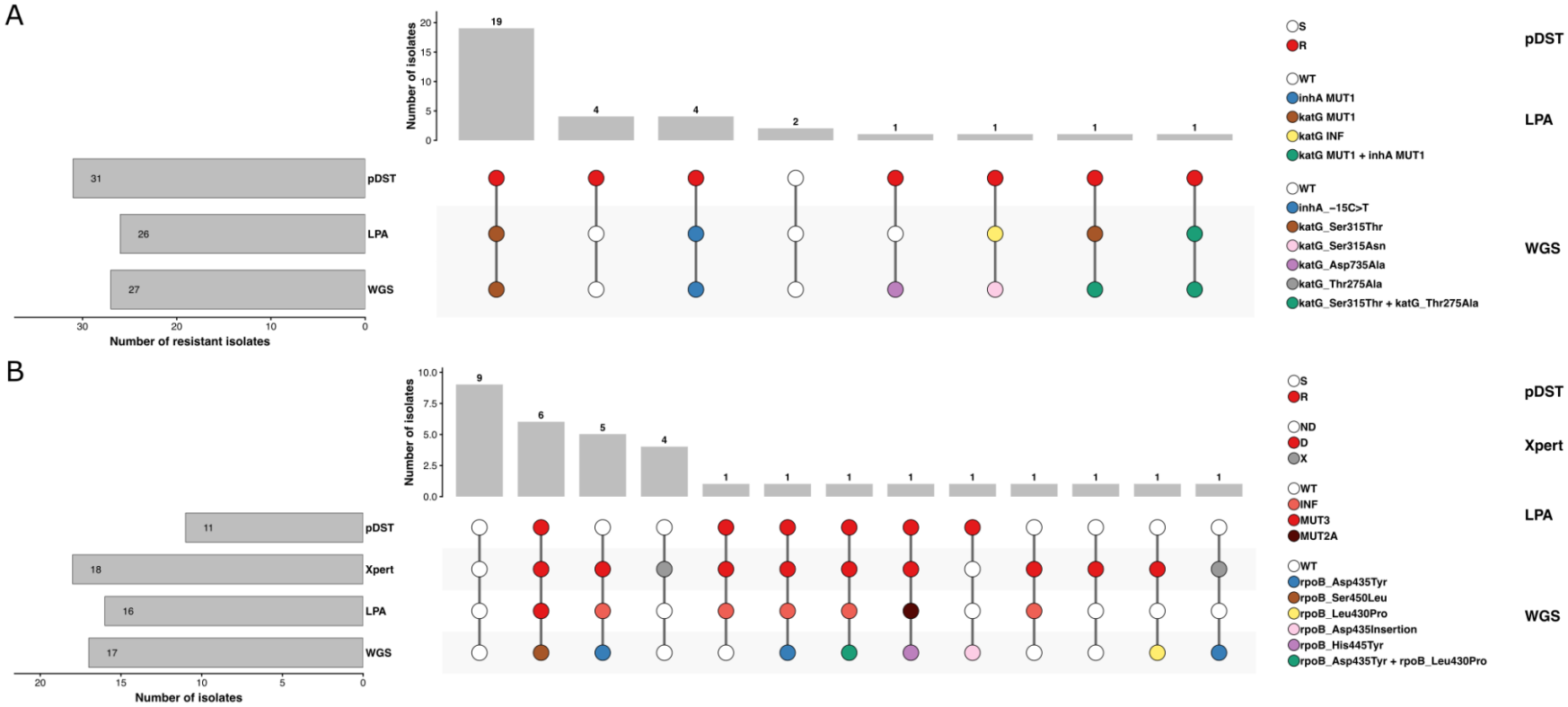
Drug resistance profiles among 33 clinical *M. tuberculosis* isolates for isoniazid (A) and rifampicin (B), assessed by phenotypic drug susceptibility testing (pDST), Xpert MTB/RIF Ultra (rifampicin only), GenoType MTBDRplus line-probe assay (LPA), and whole-genome sequencing (WGS). The UpSet plots show the distribution and concordance of resistance determinants across detection methods. For each panel, the upper bar chart shows the number of isolates sharing each unique combination of results, with counts labeled above each bar. The central matrix connects results within each profile; filled circles represent the categorical outcome for each assay, with colors corresponding to specific mutations or phenotypic classifications. White circles denote wild-type (susceptible) results. The right-side horizontal bar chart shows the total number of isolates each method classified as resistant. pDST: phenotypic drug susceptibility testing; Xpert: Xpert MTB/RIF Ultra; LPA: GenoType MTBDRplus line probe assay; WGS: whole-genome sequencing; S: susceptible; R: resistant; INF: inferred, that refers to the absence of the corresponding wild-type (WT) probe without hybridization to a specific mutant probe.

Two isolates (2/33; 6.06%) were fluoroquinolone-resistant by both pDST and LPA, with *gyrA* Asp94Asn and *gyrA* Ser91Pro confirmed by WGS, both were also resistant to rifampicin and isoniazid. One isolate from a newly diagnosed TB patient was phenotypically resistant to both bedaquiline and isoniazid (1/33; 3.03%), although WGS identified no known bedaquiline resistance-associated mutation. The complete pDST, gDST, and wgsDST data are available in **Supplementary Table 2**.

All 33 sequenced isolates belonged to Lineage 4. The most prevalent sublineages were 4.1.2 (13/33; 39.39%), 4.1.2.1 (6/33; 18.18%), and 4.3.3 (6/33; 18.18%), followed by 4.3.2, 4.3.4.1, and 4.3.4.2 (2/33 each; 6.06%) and 4.8 and 4.4.1.1 (1/33 each; 3.03%), the latter two resistant exclusively to isoniazid and detected only by pDST. Among the 27 isolates with isoniazid resistance-associated mutations, *katG* Ser315Thr predominated (21/27; 77.78%), followed by the *fabG1–inhA* operon -15C>T promoter mutation (4/27; 14.81%), with *katG* Ser315Asn and Asp735Ala in one isolate each. Rifampicin resistance-associated mutations were identified in 17 isolates (51.5%), most frequently the WHO-defined borderline mutation *rpoB* Asp435Tyr (8/17; 47.06%; one also harboring Leu430Pro) and *rpoB* Ser450Leu (6/17; 35.30%); *rpoB* Leu430Pro alone, *rpoB* His445Tyr, and a 12-nucleotide insertion at codon 435 of *rpoB* occurred in one isolate each (1/17; 5.88%). Streptomycin resistance-associated mutations were identified in 10/33 isolates (30.30%), most frequently *gid* 351delG (8/10; 80.0%), with *gid* Pro84Leu and a premature stop-gain mutation (*gid* Gln127*) in one isolate each. Ethambutol resistance-associated mutations were found in 4/33 isolates (12.12%), mainly *embB* Met306Val (3/4; 75%), with one *embA* -12C>T promoter mutation. Fluoroquinolone resistance-associated mutations were found in 2/33 isolates (6.06%), both in *gyrA*.

We next analyzed the 33 genomes sequenced in this study together with 267 genomes collected between 2011 and 2019, comprising previously published data^20,26^ and unpublished sequences generated by our research group, totalling 300 genomes from 48 municipalities collected between 2011 and 2024. Most isolates originated from the state capital, Porto Alegre (n=107), followed by the metropolitan municipalities of Alvorada (n=28), Canoas (n=19), Viamão (n=17), and Cachoeirinha (n=11). A total of 181 of the 300 isolates (60.33%) were grouped into 32 genomic clusters. We defined persistence as the presence within the same cluster of at least one isolate from the initial surveillance period (2011–2014) and at least one from a later period (2018-2024), indicating continuous circulation. Twelve clusters (37.50%) met this criterion, demonstrating sustained transmission throughout the study period (**Figure 2A**).

**Figure 2.**
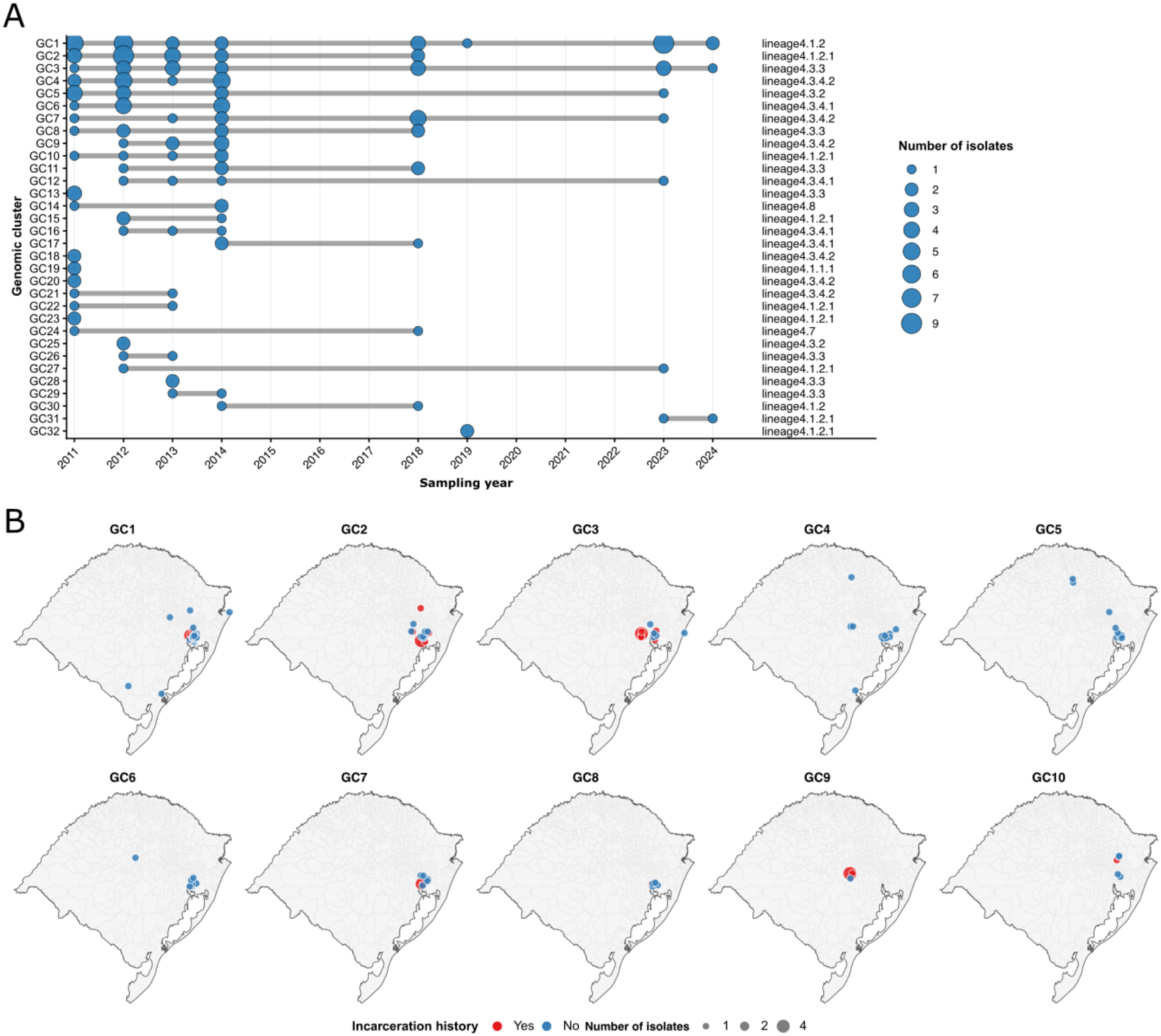
Spatiotemporal dynamics and transmission persistence of *M*.*tuberculosis* genomic clusters in Rio Grande do Sul (2011–2024). (A) Temporal distribution of 32 genomic clusters (GC1–GC32) identified among 300 clinical *M. tuberculosis* isolates over a 14-year sampling window. Each row represents an individual genomic cluster, and circles denote detected cases in a given year, with circle size scaling proportionally to the number of isolates. (B) Spatial distribution of isolates belonging to the top 10 largest genomic clusters (GC1–GC10) across municipalities of Rio Grande do Sul. Points are colored according to incarceration history. Circle area scales proportionally to the number of coincident isolates at the same location at the neighborhood level.

Of the 300 isolates, 133 (44.33%) were from newly diagnosed patients with no previous anti-TB treatment, accounting for 43.70% (52/119) of non-clustered and 44.75% (81/181) of clustered isolates, with no significant difference (p = 0.906; OR = 1.04, 95% CI 0.64–1.71). Porto Alegre accounted for the largest proportion of clustered isolates (33.15%), followed by Alvorada (12.71%), Canoas (8.29%), and Viamão (6.08%), with substantial clusters also in the regional centers of Caxias do Sul (5.52%) and Santa Cruz do Sul (4.97%).

The largest cluster (GC1) comprised 32 isolates of sublineage 4.1.2, including 10 from the current study; 26 (81.25%) originated from Porto Alegre and its metropolitan region and six were from incarcerated individuals, all collected from 2018 onwards. The second-largest cluster (GC2) comprised 20 isolates of sublineage 4.1.2.1, 16 (80%) from Porto Alegre and its metropolitan region; nine were from incarcerated individuals, whereas all isolates collected from 2013 onward were from community settings, indicating a temporal shift. Two GC2 isolates from 2018 were pre-XDR.

GC3 comprised 16 isolates of sublineage 4.3.3 RD115, 15 assigned to spoligotype SIT 863/PINI2. Eleven harbored a 12-nucleotide insertion at codon 435 of *rpoB*, identified in 2011, 2018, and 2023, indicating its persistence within the cluster; isolates without the insertion collected in 2023 and 2024 fell within the same cluster, indicating co-circulation and ongoing transmission of both variants. Most isolates were from incarcerated individuals in different prison establishments, particularly in Porto Alegre and Charqueadas. GC4, GC5, and GC6 comprised 13, 10, and 9 isolates, respectively, all from community settings, whereas smaller clusters varied in their association with incarceration: four of nine isolates in GC7, one of seven in GC8, five of six in GC9 (the remaining isolate from a person experiencing homelessness), and one of five in GC10 (**Figure 2B**). All six GC9 isolates originated from Santa Cruz do Sul. Complete information on each genomic cluster is provided in **Supplementary Table 3**.

We found no significant temporal change in sublineage distribution over the 14-year surveillance period (p = 0.144), indicating stable relative frequencies of the main circulating sublineages. In contrast, sublineage distribution differed significantly among the ten municipalities with the most sampled isolates (p < 0.001). Post-hoc standardized residuals indicated enrichment of sublineage 4.3.4.2 in Santa Cruz do Sul (*z* = 6.13) and 4.1.1.1 in Viamão (*z* = 6.03), both of which remained significant under the more conservative Bonferroni correction, together with 4.7 in Cachoeirinha (*z* = 3.67), 4.1.2 in Alvorada (*z* = 3.35), and 4.3.3 in Caxias do Sul (*z* = 3.14). No municipality showed a significant depletion of any sublineage, and Porto Alegre, Canoas, São Leopoldo, Gravataí, and Charqueadas showed no significant enrichment, consistent with a more heterogeneous distribution of circulating strains (**Figure 3**).

**Figure 3.**
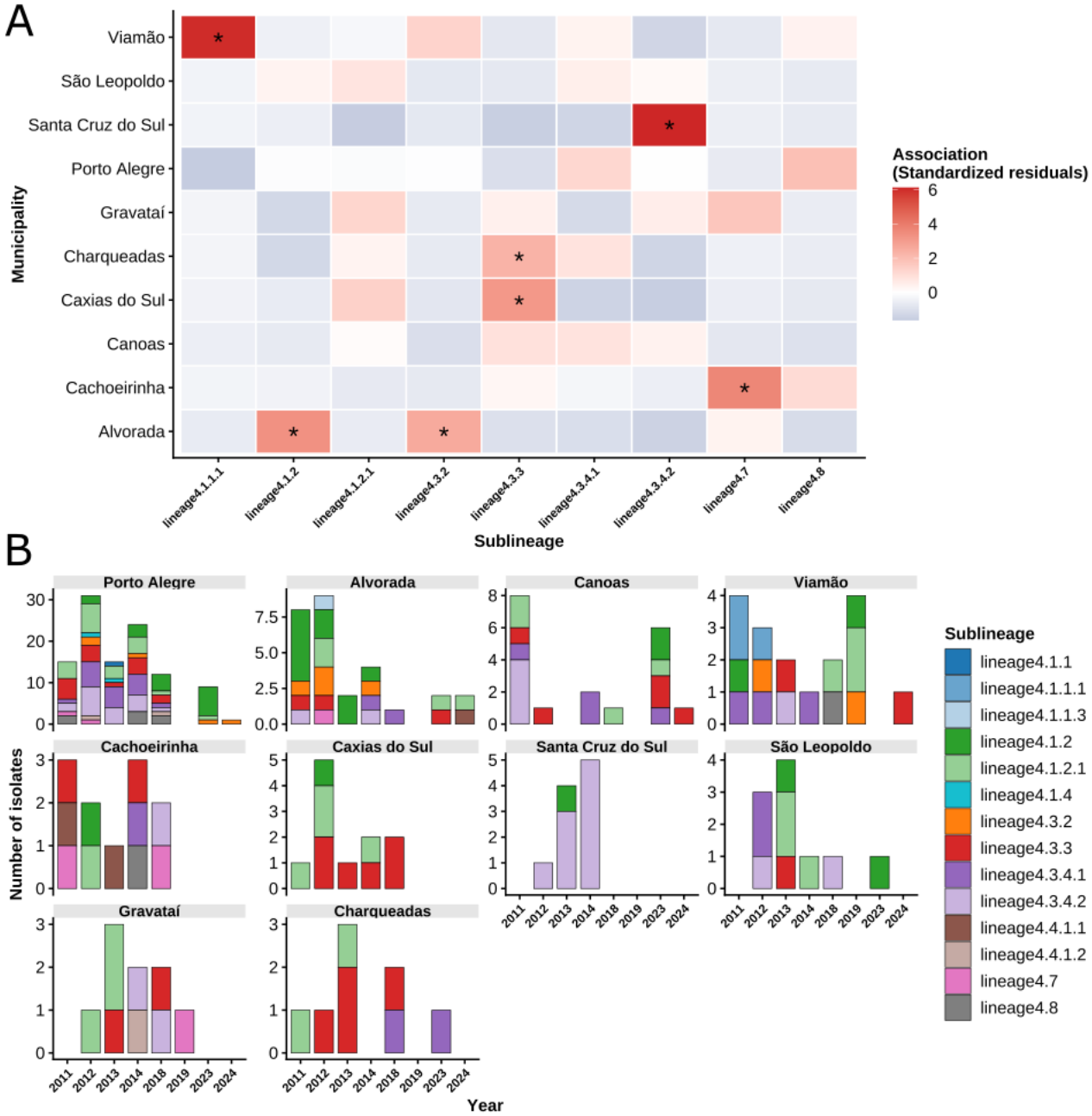
Temporal and spatial distribution of *M. tuberculosis* sublineages across municipalities in Rio Grande do Sul, Brazil, 2011–2024. **(A)** Temporal evolution of sublineages by municipality. The stacked bar chart shows the yearly distribution of genomic sublineages among the ten municipalities with the most included isolates. **(B)** Spatial association between sublineages and municipalities. The heatmap shows the standardized residuals from a chi-square test of independence between the same ten municipalities and the most frequent genomic sublineages (each present in ≥5 isolates overall). Each cell represents the association strength between a given municipality (rows) and a specific sublineage (columns), with colors indicating how much the observed counts deviate from independence. Positive residuals (red shades) indicate that the observed number of isolates from that sublineage in that municipality is higher than expected by chance (positive association). Negative residuals (blue shades) indicate that the observed count is lower than expected (negative association). White cells denote no significant deviation from the expected frequency. Asterisks (*) mark statistically significant associations at the *p* < 0.05 level (standardized residual ≥ 1.96).

Within-municipality diversity (Simpson’s Diversity Index, 1 − D) further highlighted marked spatial heterogeneity. Viamão (0.85), Porto Alegre (0.84), and Cachoeirinha (0.84) exhibited the highest diversity, whereas Santa Cruz do Sul showed extreme monoclonality (1 - D = 0.18), with 90% (9/10) of isolates belonging to sublineage 4.3.4.2, indicating localized clonal expansion. These nine isolates were grouped into two distinct clusters: GC9, linking five incarcerated individuals and one person experiencing homelessness, and GC4, grouping three residents of the same neighborhood.

Among the 300 genomes, 178 were classified as rifampicin-resistant based on resistance-associated *rpoB* mutations. Of these, 80 (44.94%) were from patients receiving their first anti-TB treatment, of which 59 (73.75%) were clustered. The remaining 98 (55.06%) were from previously treated patients (61, 62.24%, treatment failures; 37, 37.76%, losses to follow-up), of which 74 (75.51%) were clustered. Because resistance profiles at first diagnosis were unavailable for previously treated patients, the origin of rifampicin resistance in this group was assessed using the phylogenetic approach^25^. At the observed sampling fraction, transmission accounted for 88.2% (95% CI, 83.8–91.9%) of RR-TB under the N approach and 79.2% (77.2–85.4%) under the N−1 approach, with *de novo* acquisition accounting for 11.8% (8.1–16.2%) and 20.8% (14.6–22.8%), respectively (**Figure 4**), giving a plausible range of 79.2–88.2% for transmission. Extrapolating to complete sampling with SIMEX, transmission accounted for 95.2% (80.6–99.0%) and 94.7% (70.2–99.3%) under the N and N−1 approaches, respectively.

**Figure 4.**
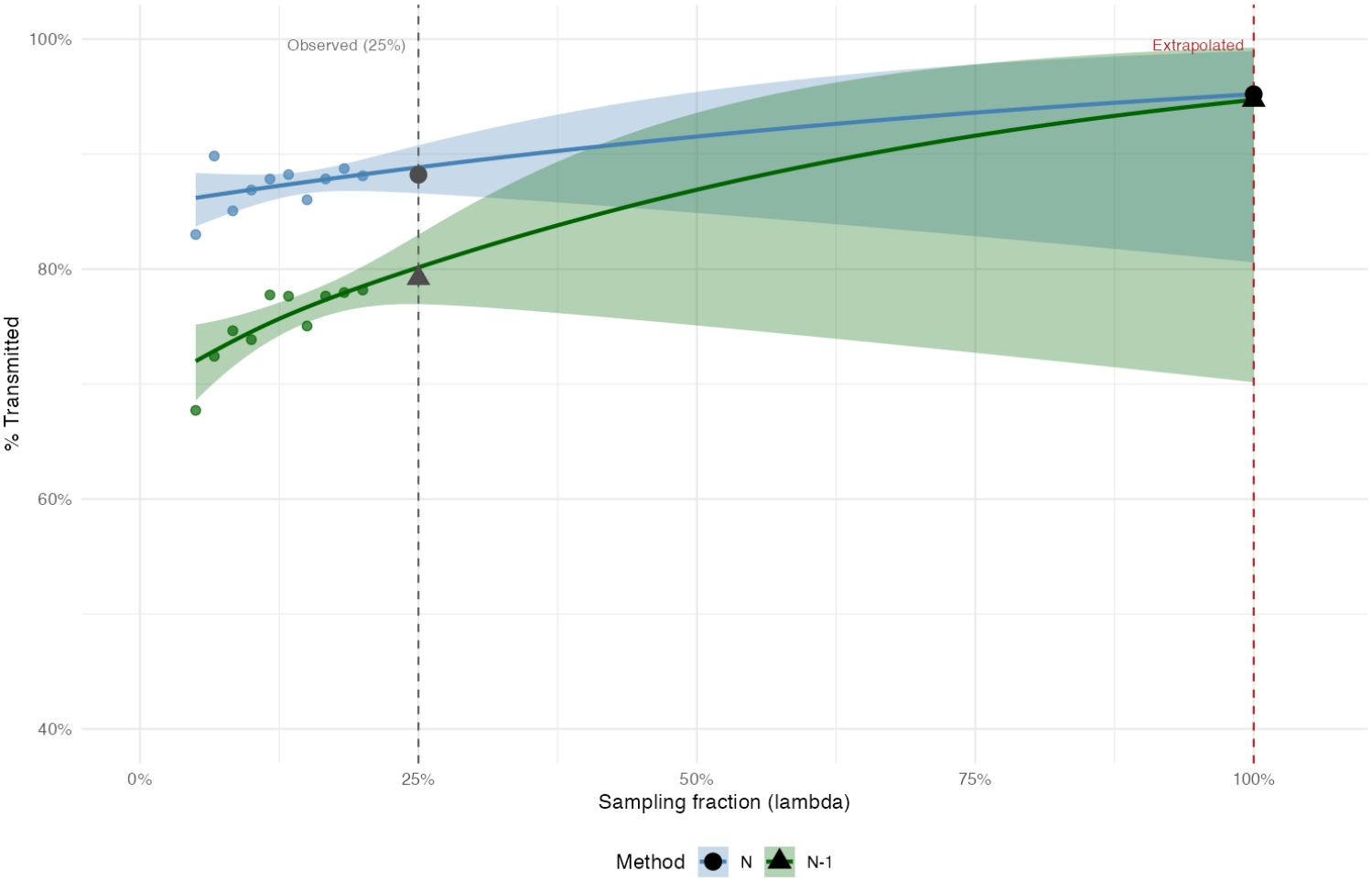
SIMEX extrapolation of transmitted rifampicin resistance under N and N−1 classification approaches. The proportion of rifampicin resistance attributable to transmission is shown across simulated sampling fractions, with the 178 rifampicin-resistant isolates as the denominator. Points represent the mean estimates obtained from repeated subsampling (five replicates per fraction), lines represent binomial generalized additive model (GAM) fits and shaded areas indicate the corresponding 95% confidence intervals. The observed estimate corresponds to the sampling fraction of 25%, whereas the extrapolated estimate at λ = 1.0 represents the expected proportion under complete sampling. The N approach provides the lower bound for de novo resistance acquisition, whereas the N−1 approach provides the upper bound. The width of the N−1 confidence interval at λ = 1.0 reflects the limited information available for extrapolation beyond the simulated range.

## DISCUSSION

Rio Grande do Sul, in southern Brazil, is among the five Brazilian states with the highest number of DR-TB cases, making it a priority setting for TB care and transmission control. By integrating WGS with epidemiological and phenotypic data, we characterized the genomic determinants of resistance and demonstrated that sustained transmission of resistant strains is the primary driver of the current RR-TB burden.

Our molecular-based resistance prediction demonstrated the importance of molecular assays for detecting first- and second-line drug resistance, particularly rifampicin resistance caused by borderline *rpoB* mutations that pDST often misses: approximately 41% of isolates classified as rifampicin-susceptible by pDST were potentially misclassified, most harboring these mutations. These confer clinically relevant resistance comparable to the canonical Ser450Leu substitution, despite reduced *in vitro* growth that may lead to false susceptibility in pDST^8,9^. Emerging variants harboring unusual resistance-conferring mutations can also be missed by molecular tools; the 12-nucleotide *rpoB* insertion, detected in one isolate here, belongs to an emerging endemic strain associated with an important genomic cluster in the state^20,26^. These findings support Brazil’s integrated approach to drug resistance diagnosis, in which LPAs, introduced into the SUS in 2021, complement pDST, which remains important for detecting resistance that molecular assays may miss^27^.

Our findings also underscore the value of molecular testing directly from sputum: Xpert MTB/RIF Ultra alone identified two isolates as rifampicin-resistant, and WGS identified heteroresistance in one, enabling earlier and more accurate treatment decisions^28^. Because the frequency of borderline *rpoB* mutations and other emerging variants varies across regions, WGS-based surveillance is essential to inform diagnostic algorithms and treatment policies in TB programs. Conversely, we identified by pDST one case of resistance to bedaquiline, recently introduced in Brazil, that WGS did not detect, in a newly diagnosed patient with no previous exposure to the drug, indicating baseline resistance. Baseline bedaquiline resistance appears to vary geographically, and its mechanisms remain largely unclear^29^; surveillance combining pDST and WGS is therefore essential at the regional level to preserve the effectiveness of this drug^30^.

Clustering analysis of 300 drug-resistant isolates collected between 2011 and 2024 revealed a clustering rate of 60.33% across 32 genomic clusters, most concentrated in the Porto Alegre metropolitan area. The persistence of several of these clusters, which likely mirror transmission chains, indicates they remained active and uncontrolled for more than a decade in both prison and community settings, with evidence of dispersal from the capital toward the state interior.

Sublineage composition remained temporally stable over the 14 years, with sustained circulation of multiple established sublineages. The marked geographic structuring observed among municipalities suggests that spatial processes played a greater role than temporal turnover in shaping population structure, consistent with localized transmission dynamics and heterogeneous population connectivity. The high diversity in metropolitan municipalities likely reflects greater population mobility and connectivity. Conversely, the predominance of sublineage 4.3.4.2 in Santa Cruz do Sul, where 90% of sampled isolates belonged to this sublineage, suggests localized expansion of a restricted genetic background; their identification in two independent genomic clusters indicates sustained circulation of different, closely related transmission chains rather than a single chain. In contrast, GC4, GC5, and GC6 comprised exclusively community-associated isolates, mostly from Porto Alegre and its metropolitan region but with genetically related isolates in other regions of the state, suggesting that some transmission chains extend beyond their main areas of concentration.

The genomic clusters identified further illustrate how transmission may be structured by social and epidemiological settings. GC1 was predominantly community-associated, whereas GC2 and GC3 showed substantial representation of isolates from incarcerated individuals, highlighting prisons as important environments for maintaining and amplifying transmission chains^31^. The predominance of prison-associated isolates in more recent years in GC1 and the shift from prison- to community-associated isolates in GC2 indicate transmission connectivity between prison facilities and the wider community. In GC3, isolates carrying and lacking the *rpoB* codon 435 insertion co-circulated across sampling years, demonstrating persistence within a sustained transmission chain rather than recent emergence of a single resistant genotype^26^.

Taken together, these findings indicate sustained transmission of *M. tuberculosis* across both prison and community settings, showing that potentially uncontrolled transmission is not restricted to prisons. Consistently, transmission was the predominant determinant of RR-TB in our analyses: in addition to the substantial proportion of RR-TB cases among patients with no previous history of TB treatment, phylogenetic-based inference attributed 79.2–88.2% of RR-TB cases to transmission. Similar estimates have been reported in Viet Nam (72–87%) using the same approach^25^, and in clustering analyses from South Africa (approximately 70%) and China (approximately 61%)^32,33^.

Several operational indicators from the TB prevention and care program may help explain the sustained transmission observed. In 2024, only 43.5% of newly diagnosed pulmonary TB cases in the state underwent rapid molecular testing (64% in Porto Alegre), culture was performed in 23% of new cases, and drug susceptibility testing in only 56% of retreatment cases with positive cultures; about 65% of identified contacts were examined, with no specific data for contacts of DR-TB cases. These indicators broadly follow the national pattern and reflect broader operational and structural challenges faced by TB programs^34^, including a high number of diagnosed cases, budgetary constraints, and laboratory capacity centralized in a small number of reference or regional laboratories. Although they do not establish a causal relationship with the transmission patterns observed, they highlight gaps in diagnostic coverage, resistance detection, and contact investigation that may facilitate the continued circulation of drug-resistant strains in both community and prison settings.

Our study has important limitations. We sequenced only a subset (52.9%; n = 36) of the 68 DR-TB isolates identified during the study period, with 33 genomes retained after quality filtering, which may not fully capture the genomic diversity of circulating strains and may have missed rare or emerging resistance variants. Furthermore, the historical dataset did not have homogeneous representation of resistant cases across the state over time, representing approximately 25% of registered cases during the period. Despite implementing the TB-SIMEX framework and simulation-extrapolation, limited sampling may still have introduced bias, as the method assumes that unsampled cases are missing at random.

In conclusion, the DR-TB burden in Rio Grande do Sul appears to result largely from uncontrolled transmission, and we found evidence of baseline resistance to the recently introduced bedaquiline, threatening the sustainability of this drug. These findings point to an urgent need for population-level interventions that prioritize resistance detection among new cases through molecular and phenotypic testing, improve contact investigation, and target persistent transmission chains, particularly in prison and community settings.

## Contributors

TSS and RSS drafted the first manuscript. AFP, FF, DBP, FG, SM, SD, GLB contributed to methodology and data generation. KW, JP, IR, PEAS and RSS contributed to critical revision of the manuscript. PEAS and RSS contributed to conceptualization, project administration, and resources. TSS and RSS accessed and verified the underlying data. All authors had access to the data and accepted responsibility for the decision to submit for publication.

## Data Availability

The raw FASTQ reads newly generated in this study have been deposited in the NCBI Sequence Read Archive (SRA) under BioProject accession PRJNA1556173. Previously published sequencing data are available under BioProject accessions PRJNA692642, PRJNA639713, and PRJNA535343. All scripts used for the SIMEX analysis are publicly available at https://github.com/salvatolab/TB-transmission-Rio-Grande-do-Sul.

https://github.com/salvatolab/TB-transmission-Rio-Grande-do-Sul

## Acknowledgements

This study was partially supported by the National Institute of Science and Technology in Tuberculosis (INCT-TB), Brazil [CNPq - 408438/2024-0]; Research Support Foundation of the State of Rio Grande do Sul (FAPERGS), Brazil [EU-LAC 25/2551-0003038-2]; and by the Coordenação de Aperfeiçoamento de Pessoal de Nível Superior (CAPES), Brazil [Finance Code 001 CAPES].

We acknowledge the support of the Brazilian Ministry of Health for providing supplies, and thank the team at the Centro de Referência Professor Hélio Fraga for conducting phenotypic drug susceptibility testing for second-line drugs. During the preparation of this work, the authors used Claude AI in order to improve language and readability. After using this tool, the authors reviewed and edited the content as needed and takes full responsibility for the content of the published article.

