## Supplementary Methods for "Long-term transmission chains fueling the drug-resistant tuberculosis burden in Southern Brazil"

#### Estimation of transmitted versus *de novo* acquired rifampicin resistance

This document gives the full specification of the TB-SIMEX analysis summarised in the main text: input construction, classification rules, simulation design, extrapolation model, uncertainty quantification, and the parameter values and software versions needed to reproduce the results.

---

#### S1. Overview

Rifampicin resistance in an individual patient may be inherited from the infecting strain (transmitted resistance) or may arise during treatment (*de novo* acquisition). The two cannot be distinguished from the genotype alone. We combined each patient's treatment history with the phylogenetic position of the resistance mutation, following the framework of Spies et al., and corrected for incomplete genomic sampling using simulation–extrapolation (SIMEX).

The analysis proceeds in six stages:

1. construction of a SNP table and variable-site alignment from the per-isolate variant calls;
2. definition of the rifampicin-resistant case set from each isolate's genotype;
3. phylogenetic reconstruction, for the complete cohort and for each simulated subsample;
4. parsimony mapping of resistance mutations onto each phylogeny;
5. classification of each resistant case as transmitted or acquired, under two bounding rules;
6. extrapolation to complete sampling, and quantification of uncertainty by bootstrap.

---

#### S2. Construction of the SNP table and alignment

Variant calls for each isolate were combined into a single SNP table with one row per variable position and one column per isolate. Every cell carries an explicit nucleotide. At positions where an isolate matches the reference, the H37Rv reference base is written out rather than being left as a gap or a dot. This is required by the mapping step: SNPPar determines whether a site is variable from the set of A/C/G/T characters observed at that position, and a gap is not a character state. A table in which reference-matching cells are encoded as gaps therefore appears to SNPPar to contain a single observed base at most sites, and those sites are silently discarded.

Filters were applied in the following order:

| Order | Filter | Rationale |
|---|---|---|
| 1 | single-base REF and ALT only | excludes indels and multiallelic records, which have no single-character state |
| 2 | PE/PPE mask | repetitive, poorly mappable regions |
| 3 | REF must match the reference genome | guards against coordinate or reference mismatches |
| 4 | DP ≥ 10 and GQ ≥ 20 | minimum depth and genotype quality |

Positions retaining fewer than two distinct bases across the cohort were dropped, since they are uninformative for both tree reconstruction and parsimony mapping. This procedure yielded 11,032 variable positions across the 300 isolates.

The same set of positions, in the same order, was written as a FASTA alignment so that the tree and the mutation mapping refer to identical sites.

---

#### S3. Definition of the resistant case set and of the denominator

Isolates were classified as rifampicin-resistant on the basis of their own genotype: any resistance-associated *rpoB* mutation reported by TB-Profiler. Of the 300 isolates, 178 (59.3%) met this definition.

The denominator for all proportions is this set of 178 cases. It is defined diagnostically and is independent of the phylogeny: a susceptible isolate nested within a clade of resistant isolates is never counted, and the denominator does not change with tree topology or with sampling fraction.

Within the 178, a subset carries a mutation that parsimony can place on the tree:

| Category | n | Treatment in the analysis |
|---|---|---|
| Rifampicin-resistant (any *rpoB* mutation) | 178 | denominator |

| Category | n | Treatment in the analysis |
|---|---|---|
| …carrying a non-synonymous RRDR SNV | 169 | eligible for mapping |
| …with an event actually mapped on the observed tree | 164 (92.1%) | classified from the tree |
| …without a mappable event | 14 | counted as transmitted by default |

The 14 cases without a mappable event are of two kinds. Twelve carry only an indel: eleven the 12-bp in-frame insertion c.1304delAinsGCCAGAACAACCC, and one the 6-bp deletion c.1300_1305delATGGAC. The remaining two carry an RRDR SNV at sub-fixed frequency (p.Leu430Pro at 0.55 and p.His445Asp at 0.33). Neither an indel nor a minority-frequency allele produces a fixed single-nucleotide state that can be reconstructed by parsimony, so no event is placed for these isolates.

For the eleven isolates carrying the 12-bp insertion, classification as transmitted is supported by the phylogeny rather than assumed. All eleven fall within a single clade; the insertion is therefore shared by descent from a common ancestor and was not acquired independently on eleven occasions.

The default classification bears only on previously treated patients, since treatment-naive patients are classified as transmitted on clinical grounds alone.

---

### S4. Phylogenetic reconstruction

Maximum-likelihood phylogenies were reconstructed with IQ-TREE v3.0.1 under TVM+F+ASC, the model selected by ModelFinder for the complete cohort, with an ascertainment-bias correction appropriate to an alignment of variable sites.

The same procedure, model and site filtering were applied to the complete cohort and to every simulated subsample. This matters because the SIMEX trend is a comparison between the observed estimate and the simulated ones: if the observed tree were built under a different model or from a differently filtered alignment, part of any apparent trend would reflect that difference rather than sampling fraction.

Sites invariant within a given subsample were removed from that subsample before reconstruction. Two reasons: the ascertainment-bias correction assumes an alignment containing no constant sites, and retaining them would let the number of informative sites grow with the sampling fraction, confounding the quantity the SIMEX model is fitting.

#### S5. Rooting

All trees were rooted on a single common reference group, defined once from the smaller side of the root bipartition of the complete-cohort tree and then applied to every replicate.

Rooting each replicate independently — by midpoint, for example — would place topologically different roots in different subsamples. Since the classification depends on the inferred direction of mutation along each branch, independent rooting would introduce variation in the estimate that is an artefact of the rooting procedure rather than of sampling fraction. A replicate retaining no member of the reference group falls back to midpoint rooting; in the analysis reported here no replicate required this fallback.

#### S6. Mapping of resistance mutations

Mutations were mapped onto each rooted phylogeny by maximum parsimony using SNPPar v1.0, with the H37Rv (NC_000962.3) GenBank record for annotation.

For each run, the master SNP table was reduced to the columns of that run's taxa, and rows no longer showing at least two distinct bases were dropped, so that the table given to SNPPar is consistent with the alignment from which the tree was built.

Events were restricted to non-synonymous changes in *rpoB* (Rv0667) at the codons of the rifampicin resistance-determining region: 170, 426–452 and 491, in *M. tuberculosis* numbering.

SNPPar assigns each event either to a tip, meaning the mutation is unique to that isolate, or to an internal node, meaning it is shared by all descendants of that node and therefore arose once in their common ancestor.

#### S7. Classification rules

Patients with no history of previous TB treatment were classified as having transmitted resistance, since the absence of prior drug exposure precludes treatment-mediated selection.

For patients with a history of previous treatment, classification follows the phylogenetic position of the resistance mutation.

Terminal events. A resistance mutation on a terminal branch arose in that patient's own infection. In a previously treated patient, this is a *de novo* acquisition.

Internal-node events. Where a resistance mutation maps to an internal node, resistance arose once in the common ancestor of the descendants, but which descendant lineage experienced the acquisition cannot be identified. Two bounding rules are therefore applied:

- N approach — all descendants of the internal resistance node are counted as transmitted. This is the lower bound for *de novo* acquisition.

- N−1 approach — exactly one *de novo* acquisition is assigned per such node, when at least one descendant patient had a history of previous TB treatment. The remaining descendants are counted as transmitted. This is the upper bound.

Two details of the N−1 rule are worth stating explicitly, since they determine the behaviour in the cases most likely to be queried:

1. The assignment is one acquisition per node, irrespective of how many descendants were previously treated. A node with five previously treated descendants contributes the same single acquisition as a node with one. The rule bounds the number of independent acquisition events, not the number of previously treated patients.
2. A node whose descendants were all treatment-naive contributes no acquisition under either rule.

The total number of acquisitions is capped at the number of rifampicin- resistant cases, so that the estimated proportion cannot exceed 100%.

Together the two rules define the plausible range of transmitted and *de novo* acquired rifampicin resistance.

---

#### S8. SIMEX simulation design

The observed cohort corresponds to λ = 0.25, that is 25% of notified rifampicin-resistant cases in the study period were sequenced. SIMEX works by making the sampling deficiency progressively worse in a controlled way, observing how the estimate responds, and projecting that relationship back to complete sampling.

The cohort was subsampled without replacement at ten evenly spaced sampling fractions from λ = 0.05 to λ = 0.20, with five independent replicates per fraction, giving 50 simulated datasets plus the complete cohort.

This grid follows the rule implemented in the TB-SIMEX reference pipeline: evenly spaced levels from λ = 0.05 up to 0.05 below the observed sampling fraction. Expressed relative to the sequenced set, it retains 20–80% of isolates. At the 0.64 sampling coverage reported by Spies et al. the same rule corresponds to retaining between 8% and 92% of sequenced isolates, which is how the grid is described in that publication; the difference in the relative figures follows from the difference in sampling coverage, not from a different rule.

Because only subsampling downward is possible, the simulated range cannot extend above the observed sampling fraction. The consequences for interpretation are addressed in S9.

---

#### S9. Extrapolation model and diagnostics

Estimates across sampling fractions were modelled with a binomial generalized additive model, logit link, with a thin-plate regression spline on λ (bs = "tp", k = 10), fitted by REML, and predicted at λ = 1.0.

Because the simulated range reaches only λ = 0.20 while the prediction target is λ = 1.0, the extrapolation projects roughly four times beyond the data. The estimate at the observed sampling fraction is therefore reported as the primary result, and the extrapolated value is reported as a sensitivity analysis, accompanied by three diagnostics that indicate how much information supports it:

| Diagnostic | What it indicates |
|---|---|
| significance of the linear trend in λ | whether the estimate changes detectably with sampling fraction at all |
| effective degrees of freedom of the spline | whether the fitted curve has any curvature, or has collapsed to a straight line |
| quasibinomial dispersion (ϕ) | whether variation across replicates matches the binomial expectation |

In the analysis reported here the trend was not statistically significant under the N approach ($p = 0.19$), the spline collapsed to a linear term (edf = 1.0), and dispersion was below the binomial expectation (ϕ = 0.39–0.79). These diagnostics are the reason the extrapolated value is presented with its full confidence interval and not as a point estimate.

---

#### S10. Bootstrap

Uncertainty at the observed sampling fraction was quantified by a 100- iteration bootstrap that propagates both sources of variation relevant here: which patients entered the cohort, and which phylogeny was inferred from them.

Each iteration proceeds as follows:

1. resample the cohort of 300 isolates with replacement;
2. write an alignment of the unique isolates drawn, retaining each isolate's multiplicity as a resampling weight — a phylogeny cannot be built from duplicated sequences;
3. reconstruct the phylogeny *de novo* and root it on the common reference group;
4. remap resistance mutations with SNPPar;
5. recompute the N and N−1 classifications, with each isolate contributing its resampling weight to both numerator and denominator.

Drawing *n* isolates with replacement from *n* leaves about 63% distinct, so each replicate tree carries roughly two thirds of the cohort; the median across iterations was 192 unique taxa and a weighted denominator of 180.

Confidence intervals are the 2.5th and 97.5th percentiles across the 100 iterations. They are therefore empirical and carry phylogenetic uncertainty, unlike an interval derived from the standard error of the GAM, which describes only the fitted curve.

---

#### S11. Software, parameters and random seeds

| Tool | Version | Role |
|---|---|---|
| IQ-TREE | 3.0.1 | maximum-likelihood phylogenies |
| SNPPar | 1.0 | parsimony mapping of mutations |
| R | ≥ 4.3 | subsampling, classification, GAM, figures |
| Python | ≥ 3.9 | SNP table construction |

R packages: ape, phytools, dplyr, readr, tidyr, stringr, purrr, mgcv, ggplot2, scales.

| Parameter | Value |
|---|---|
| Observed sampling fraction (λ) | 0.25 |
| Sampling-fraction grid | λ = 0.05–0.20, ten evenly spaced levels |
| Equivalent retained fraction of sequenced isolates | 20–80% |
| Replicates per sampling fraction | 5 (50 simulated datasets) |
| Bootstrap iterations | 100 |
| Substitution model | TVM+F+ASC |
| Resistance gene | Rv0667 (*rpoB*) |
| RRDR codons | 170, 426–452, 491 |
| Minimum depth / genotype quality | DP ≥ 10, GQ ≥ 20 |
| GAM | binomial, thin-plate spline on λ, k = 10, REML |

Three independent random streams were used, so that any stage can be rerun without disturbing the others: one for the SIMEX subsamples, one for the bootstrap draws, and one for the IQ-TREE searches. Seed values are given in the analysis code. Seeding IQ-TREE matters for reproducibility because an unseeded search can return a slightly different tree on reruns and shift the classification by one or two cases; with a fixed seed, trees are reproduced byte for byte on a single thread, and reproduced in topology — the quantity the classification depends on — when multiple threads are used.

The estimate at the observed sampling fraction does not depend on any random seed, since it uses the complete cohort with no subsampling.

---

#### S12. Assumptions and limitations

Treatment history is a proxy. The classification rests on recorded history of previous TB treatment. Unrecorded prior treatment would cause acquisition to be misclassified as transmission.

Resistance profiles at first diagnosis were unavailable for previously treated patients, which is why the phylogenetic approach is needed for that group in the first place.

Fourteen resistant cases have no mappable event and are counted as transmitted by default (S3). This is conservative for the *de novo* estimate.

The extrapolation has a short lever arm. The simulated range reaches $\lambda = 0.20$ against a target of $\lambda = 1.0$, a consequence of the sampling coverage achieved rather than of the grid design: no grid can extend above the observed sampling fraction, because only subsampling downward is possible. This is why the observed estimate, not the extrapolated one, is the primary result.

Parsimony mapping assumes a single origin per event where the data are compatible with one. Reversions and recurrent mutation at the same codon are reconstructed as the smallest number of changes consistent with the tree, which may not be the true history in every case.

---

### S13. Code availability

All analysis scripts, with the parameter values and random seeds used, are available at [https://github.com/salvatolab/B19V-genomic-epidemiology-Rio-Grande-do-Sul]. These scripts were adapted from the original published by Spies et al. 2026 [https://doi.org/10.1016/j.lanwpc.2026.101906], available at [https://github.com/RuanSpies21/TB-SIMEX/].
